# Spatial and hierarchical Bayesian analysis to identify factors associated with caesarean delivery use in Ethiopia: evidence from national population and health facility data

**DOI:** 10.1101/2022.10.07.22280820

**Authors:** Teketo Kassaw Tegegne, Catherine Chojenta, Theodros Getachew, Roger Smith, Deborah Loxton

## Abstract

**Background:** Caesarean section has a significant role in reducing maternal and neonatal mortality. A linked analysis of population and health facility data is valuable to map and identify caesarean section use and associated factors. This study aimed to identify geographic variation and associated factors of caesarean delivery in Ethiopia.

**Method:** Linked data analysis of the 2016 Ethiopia Demographic and Health Survey (EDHS) and the 2014 Ethiopian Service Provision Assessment Plus (ESPA+) survey was performed. Spatial analysis was conducted to identify geographic variations and factors associated with caesarean delivery. Hierarchical Bayesian analysis was also performed to identify factors associated with caesarean delivery using the SAS MCMC procedure.

**Results:** Women’s age and education, household wealth, parity, antenatal care (ANC) visits, and distance to caesarean section facility were associated with caesarean delivery use. Women who had ≥4 ANC visits were 4.67 (95% Credible Interval (CrI): 2.17, 9.43) times more likely to have caesarean delivery compared to those who had no ANC visits. Women who had education and were from rich households were also 2.80 (95% CrI: 1.83, 4.19) and 1.80 (95% CrI: 1.08, 2.84) times more likely to have caesarean deliveries relative to women who had no education and were from poor households, respectively. A one-kilometer increase in distance to a caesarean section facility was associated with an 88% reduction in the odds of caesarean delivery (Adjusted Odds Ratio (AOR) = 0.12, 95% CrI: 0.01, 0.78). Hotspots of high caesarean section rates were observed in Addis Ababa, Dire Dawa, and the Harari region. In addition, women’s age at first childbirth and ≥4 ANC visits showed significant spatially varying relations between caesarean delivery use across Ethiopia.

**Conclusion:** Caesarean section is a lifesaving procedure, and it is essential to narrow disparities to reduce maternal and neonatal mortality and avoid unnecessary procedures.

## Introduction

Caesarean section or caesarean delivery is a surgical procedure to deliver a baby, which is often performed when vaginal delivery is not possible or puts the baby and/or the mother at risk (1). A medically indicated caesarean section is effective in saving the lives of mothers and newborns (2). The World Health Organization (WHO) stated that a caesarean section rate above 10% at the population level was not associated with a reduction in maternal and newborn deaths (3, 4). A recent analysis of global trends in caesarean deliveries reported that 21.1% of deliveries were by caesarean section worldwide (5). It is also projected that by 2030, 28.5% of deliveries will be by caesarean section globally (5).

Despite an increasing trend in caesarean sections globally, inequalities exist between and within countries (5). Inequalities in caesarean sections could be explained by different demand and supply-side factors. The demand-side factors, including individual, household, and community-level factors, are significantly associated with caesarean delivery use (6-8). The supply-side factors, such as availability and quality of services, facilities’ readiness to provide services, and providers’ competency and caring behaviour, influence women’s use of caesarean sections. A systematic review reported that health facilities in many developing countries are underperforming in these areas (9).

In Ethiopia, there was a slight increase in the national rate of caesarean sections from 2000 (0.7%) to 2016 (1.9%) (10). However, there is a gap in the availability and use of caesarean sections depending on geographic locations. It is, therefore, difficult to identify areas with unmet needs and the types of interventions required to meet caesarean section needs. A linked analysis of population and health facility data has a significant role in examining the link between population healthcare needs, service utilization and the health service environment (11, 12).

In Ethiopia, the national population and health facility data can be accessed from the Ethiopia Demographic and Health Survey (EDHS) (13) and the Ethiopian Service Provision Assessment Plus (ESPA+) survey (14), respectively. Mapping caesarean delivery use and associated factors using these data would provide valuable information for program managers and decision-makers and help to identify communities requiring special attention. Thus, this study aimed to identify: 1) geographic variations in caesarean delivery use – spatial analysis, and 2) factors associated with caesarean delivery use – multilevel analysis in Ethiopia using the national population and health facility data.

## Methods

### Data sources

This study used two data sets: the 2016 EDHS and the 2014 ESPA+ survey.

#### Population data

The 2016 EDHS was used to obtain national population data. It contained population socio-demographic characteristics, health service use and morbidity/mortality data. The survey used a stratified multistage sampling technique. EDHS clusters, which were census Enumeration Areas in Ethiopia, were used as sampling frames. In addition to population information, the latitude and longitude coordinates of each EDHS clusters was recorded. Data were collected from 645 EDHS clusters (15). In this analysis, we included 6,954 women who gave birth in the five years preceding the survey from 622 EDHS clusters. We excluded 239 women who gave birth during this period from 23 EDHS clusters since they had no geographic coordinates for data linkage.

#### Health facility data

The 2014 ESPA+ survey provided national health facility data covering different aspects of the availability and readiness of health services. In addition, the data set contained the latitude and longitude coordinates of each health facility. The survey used a combination of census and simple random sampling techniques (16). Due to their importance and limited numbers, the survey included all hospitals. Details of the sampling procedure are discussed elsewhere (12). In this analysis, we included only hospitals proving caesarean sections. Thus, we included only 179 of the 214 hospitals that reported providing caesarean sections.

### Data linking method

This study used an Euclidean buffer link to link population and health facility data (12). This is regarded as the method of choice for linking a census of health facilities with population data (11, 12, 17). Details of this method are discussed elsewhere (12). The administrative boundaries of Ethiopia, obtained from Natural Earth (18), were used.

### Health service environment

Using the ESPA+ survey, four health service environment variable scores were computed. These were the average distance to the nearest caesarean delivery facilities, comprehensive obstetric care availability score, readiness to provide comprehensive obstetric care score, and a general health facilities readiness score. Using principal component analysis, availability and readiness scores were computed for the nearest hospital providing caesarean delivery. The SCORE procedure in SAS was used to compute availability and readiness scores. The comprehensive obstetric care indices were created using the World Health Organization’s ‘Service Availability and Readiness Indicators’ (19, 20). Details of computing these scores were discussed elsewhere (12).

Regarding general service readiness, six service readiness dimensions were used: communication equipment, external supervision, client opinion and feedback, quality assurance, emergency transport, and client latrine. The first two principal components (health facility management system and infrastructure) were used to compute two general service readiness scores (12).

For hospitals providing caesarean delivery, indices of comprehensive obstetric care availability and readiness scores were created. Two comprehensive obstetric care availability scores (basic and comprehensive components) were created using seven variables. Similarly, two comprehensive obstetric care readiness scores (equipment and supplies, and skilled personnel) were computed using nine dichotomous variables (12).

### Study variables

The outcome variable of this study was caesarean delivery use. A woman was considered to have used caesarean delivery if her most recent birth in the five years preceding the survey was via caesarean section.

Sociodemographic and obstetric characteristics and health facility variables were considered independent variables for this study. Since this study is a multilevel analysis, all independent variables included are of three types: level 1, level 2 and level 3. Level 1 or individual-level variables include women’s age, education, and occupation, women’s autonomy in healthcare decision-making, husband’s education and occupation, household wealth, age at first childbirth, parity, antenatal care (ANC) visits, and nature of recent pregnancy (planned or unplanned). Level 2 or EDHS cluster-level variables were general service readiness, comprehensive obstetric care availability and service readiness, and average distance to the nearest caesarean delivery facility. One level 3 or region-level (number of caesarean sections providing hospitals per region) was considered in the analysis.

Respondents’ educational status was grouped into two categories: had no education and had an education. Similarly, occupational status was grouped as having work and no work. Respondents who responded not working at the time of the interview or did not work in the last 12 months before the survey were grouped as having no work. The EDHS household wealth quintiles were also grouped into two categories: poor (including lowest, second and middle quintiles) and rich (including fourth and highest quintiles). ANC visits were grouped as no ANC visits, 1 – 3 ANC visits and ≥4 ANC visits.

### Statistical analysis

#### Bayesian hierarchical model

The EDHS survey employed a multistage cluster sampling technique: women were nested within EDHS clusters and then clusters were nested within regions. Considering this hierarchical nature of the data, a three-stage Bayesian hierarchical model was used. Women’s data within the respective EDHS clusters were also linked with the health facility data. In this model, the probability of caesarean delivery is allowed to vary both between EDHS clusters and between regions. To model the between-EDHS cluster and between-region variability, the logarithms of the odds ratios of caesarean delivery was assumed to have a normal distribution. The means of the normal distribution of log odds ratios across EDHS clusters and regions, therefore, represent the average effect in the EDHS clusters and regions, and the variances represent the variability among EDHS clusters and regions.

Bayesian inference allows for the combination of existing knowledge with new information according to established rules of probability. In this study, a non-informative prior that would not have an influence on the posterior distribution was chosen (21, 22). For each fixed effect, a non-informative prior that follows a normal distribution with large variance (*σ*^2^=1000) and a zero mean (*μ*=0) was used. Similarly, for the random effects, non-informative priors that were set up to follow a normal distribution with zero mean and different variances were used. The variances for the random effects were set up to follow an inverse gamma distribution with a shape parameter of *α*=0.01 and a scale parameter *β*=0.01. The MCMC procedure (PROC MCMC) in SAS was used to estimate the Bayesian hierarchical generalized linear mixed models. The study used a simulation size of 175,000 iterations along with 25,000 burn-in iterations. One of every 25 samples was kept, which gave the Markov chain Monte Carlo (MCMC) sample size of 7,000 for computing the posterior quantities of interest. This analysis procedure enabled the identification of potential factors associated with the rate of caesarean delivery with an equal-tail 95% credible interval.

The model building process was started with the unconditional model (a model containing no predictors) and more complex models were gradually built by checking improvements in model fit after each model is estimated (23). A Deviance Information Criterion (DIC) was calculated using the posterior mean estimates of the parameters to select the best fitting model. The unconditional (empty) model was used to calculate the intra-class correlation coefficient (ICC), which estimates how much variation in the use of caesarean delivery exists between EDHS clusters (level-2 units) and between regions (level-3 units) using posterior samples.

Three variance estimates were used to calculate EDHS cluster ICC and region ICC. The formula for calculating each of these ICCs is similar to the formula used for two-level ICC calculations. However, the denominator now has three elements (residual variance, cluster variance and region variance) which form the total variance in the model. The two equations for the EDHS cluster ICC (*ICC*_*C*_) and region ICC (*ICC*_*R*_) are below.

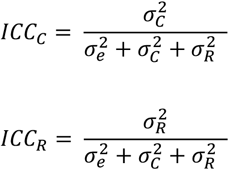

The *ICC*_*C*_ tells us how much of the total variation in caesarean delivery exists between EDHS clusters. On the other hand, *ICC*_*R*_ indicates the total variation in caesarean delivery that exists between regions (24). In hierarchical generalized linear mixed models, it is assumed that there is no level-1 error variance; however, to calculate the intra-class correlation coefficient, a slight modification is made. The level-1 residual variance (***ε*)** follows a logistic distribution and is standardized with a mean of zero and variance = *π*^2^/3 (25). Therefore, for three-level random intercept hierarchical generalized linear mixed model with an intercept variance of 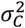 and 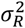, the intra-class correlation coefficient (Rho) is given by;

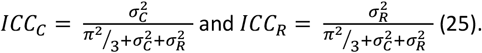

Using posterior samples from the MCMC simulation, proportional change in variance (PCV), for EDHS clusters and regions, was also estimated to measure changes in area level variance between empty and individual level models and between successive models (26, 27). This was computed using a mathematical equation: 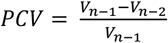; where *V*_*n*−1_ is the neighbourhood variance in the empty model and *V*_*n*−2_ is the neighbourhood variance in the subsequent model.

Based on the model fit statistics, allowing EDHS cluster-level variables to vary across regions and adding a region-level variable did not show improvement in model fit statistics. Therefore, a model with six individual-level variables (women’s age, age at first childbirth, education, household wealth, parity, and the number of ANC visits, and one EDHS cluster level variable (average distance to the nearest caesarean delivery facility) was selected. In this model, individual-level variables were allowed to vary across EDHS clusters and regions.

#### Spatial analysis

ArcGIS 10.6.1 and R version 4.2.0 were used to carry out the spatial analysis. The Ethiopian Polyconic Projected Coordinate System was used to flatten the Ethiopian map (12). Hotspot analysis and spatial regression were performed to identify spatial clusters and factors associated with spatial variations of caesarean delivery use. The unit of spatial analyses were EDHS DHS clusters. Three procedures were followed to carry out the spatial analysis as discussed elsewhere (12). First, the Global Moran’s I statistic, which is a global measure of spatial autocorrelation (28) was carried out. Second, the Incremental Spatial Autocorrelation was used to determine the critical distance at which there is maximum clustering (12). The average distance *(15 kilometres)* at which a feature has at least one neighbour was calculated. This gave 155 kilometres at which clustering of caesarean delivery use peaked. Lastly, the Getis-Ord Gi* statistic was used to identify statistically significant spatial clusters of high caesarean delivery rates (hotspots) and low caesarean delivery rates (cold spots). A False Discovery Rate (FDR) correction method was applied to account for multiple and spatial dependence tests in Local Statistics of Spatial Association (12, 29). Statistical significance was determined based on z-scores and p-values (12).

Furthermore, spatial regression was performed to identify key factors behind the observed spatial patterns of caesarean delivery use. Moran eigenvector spatially and non-spatially varying coefficient (M-SNVC) model was used to model spatially and non-spatially varying relations. Eigenvector spatial filtering regression model removes spatially autocorrelated residuals and improves model fit (30, 31). M-SNVC model is also stable and quite robust against spurious correlations compared to the Moran eigenvector spatially varying coefficient model (32). Since the spatial regression was done at the EDHS cluster level, the number of caesarean deliveries per EDHS cluster was first computed. Then we used a transformation-based generalized spatial regression using the *spmoran* package implemented in R (33). Since the count data did not obey the Poisson distribution, the log-Gaussian approximation was applied first to roughly normalize the data, and then the SAL transformation to identify the most likely distribution (33). Finally, the transformed result from the generalized spatial regression model was used in the M-SNVC model to model the spatial relationship between caesarean delivery use and explanatory variables.

### Ethics approval

This study used secondary data sets: 2016 EDHS and 2014 ESPA+ collected previously where confidentiality information was maintained. Hence, as we did not collect data directly from participants, recruitment and contacting participants were not required. Ethical approval was obtained from The University of Newcastle, the Ethiopian Public Health Institute and the Measure DHS program to access the datasets.

## Results

### Sociodemographic and obstetric characteristics

The sociodemographic and obstetric characteristics of the women were not presented in this paper. These characteristics can be found in our previous publications with different outcomes (34, 35). In this paper, women’s sociodemographic and obstetric characteristics are similar to these papers.

### Hospital characteristics

Amongst the 214 hospitals, 200 (93.46%) reported providing normal delivery care. Out of the 200 hospitals, 179 (89.50%) provided caesarean delivery. The national average distance of caesarean delivery providing hospitals to the 2016 EDHS clusters was 33.27 km. The 2016 EDHS sampled clusters in the Somali region had the longest distance to caesarean delivery providing hospitals, 71.36 km. Conversely, on average, EDHS clusters in Addis Ababa were 1.54 km away from caesarean section providing hospitals (Fig 1).

**Figure 1:**
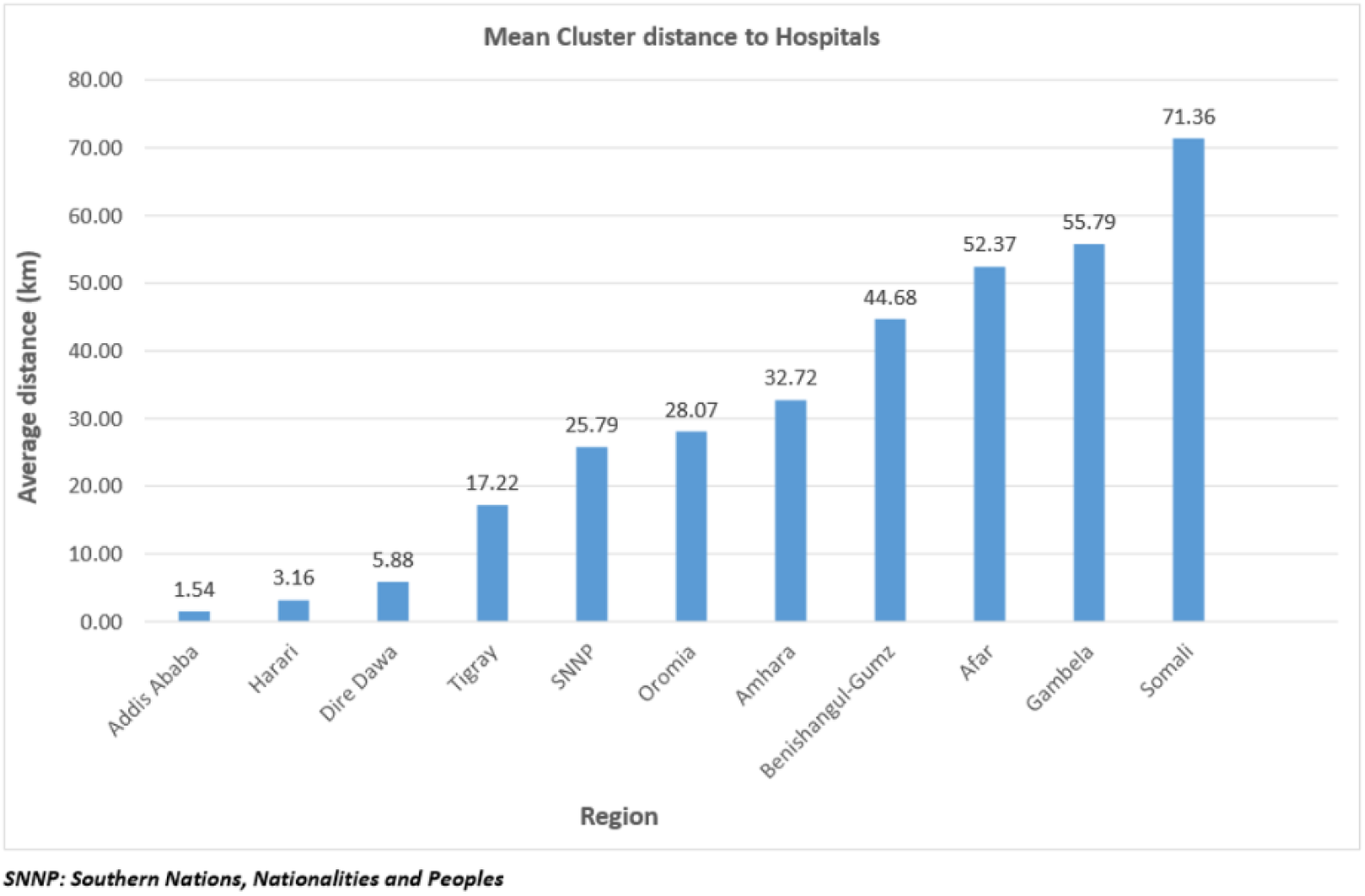
The average distance of the demographic and health survey clusters to caesarean delivery providing hospitals in Ethiopia, 2016 (n = 179)

### Prevalence of caesarean delivery

In Ethiopia, the prevalence of caesarean delivery was found to be 3.65% (urban 12.70%, 1.14% rural). The prevalence of caesarean deliveries were 7.34% in the public and 30.20% in private health facilities. The prevalence of caesarean delivery varied across the different regions and city administrations of the country; the highest caesarean delivery rate was reported in Addis Ababa (21.87%) followed by Harari (11.19%) and Dire Dawa (8.31%). The map (Fig 2) showed that there is regional variation in caesarean delivery rates.

**Figure 2:**
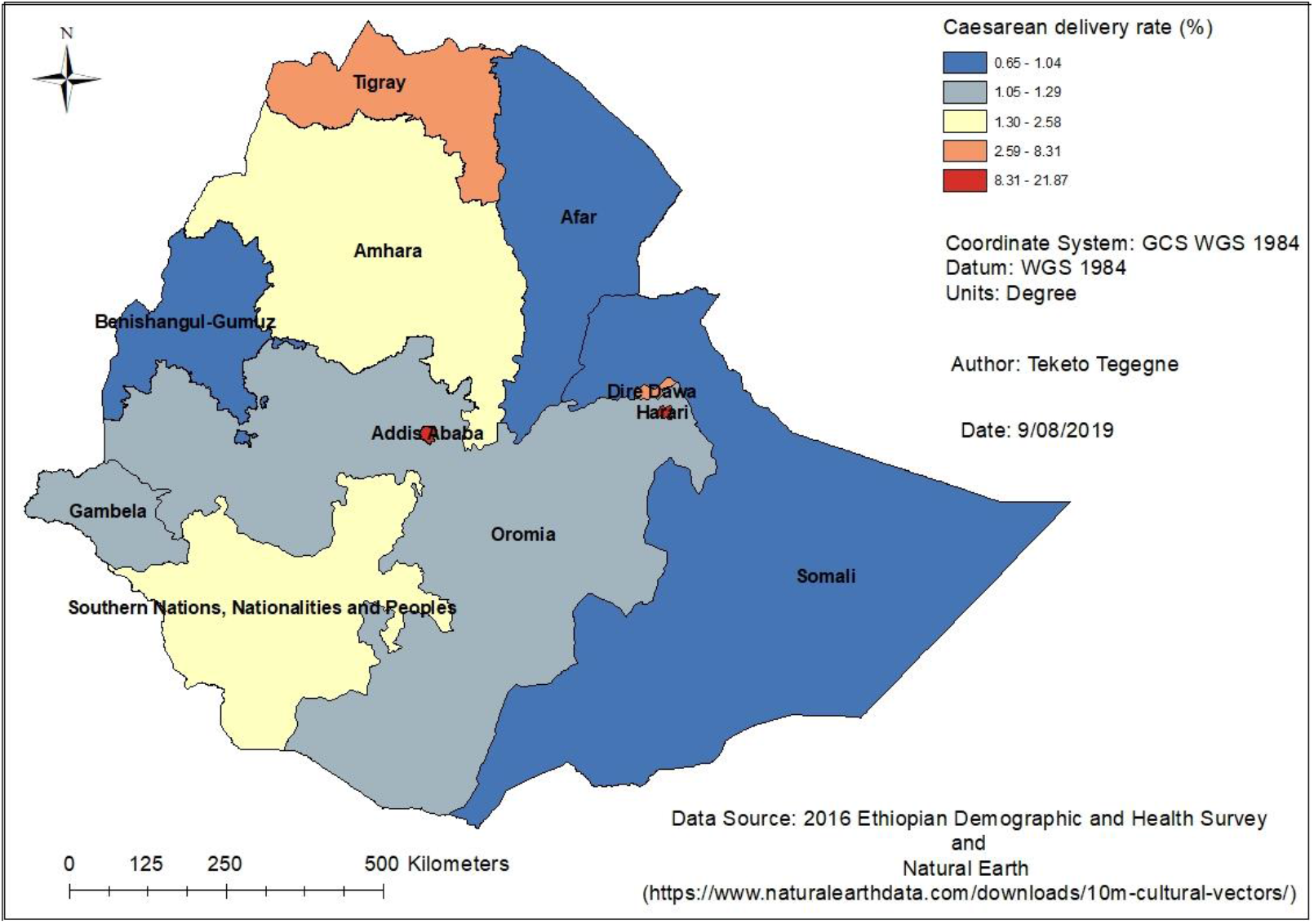
Caesarean delivery use among pregnant women in Ethiopia, 2016

### Hospital to the population proportion

There is a huge difference in the proportion of hospitals per one-million population in each region. In the Amhara, Oromia and Southern Nations, Nationalities and Peoples (SNNP) regions, approximately one hospital is expected to serve one million people. In the Tigray region, it was estimated that six hospitals serve one million people. In the two city administrations (Addis Ababa and Dire Dawa) and the Harari region, an average of 14 hospitals serve one-million people. The rates of caesarean delivery per EDHS clusters also varied depending on the number of, and distance to, caesarean delivery providing hospitals in each region (Fig 3).

**Figure 3:**
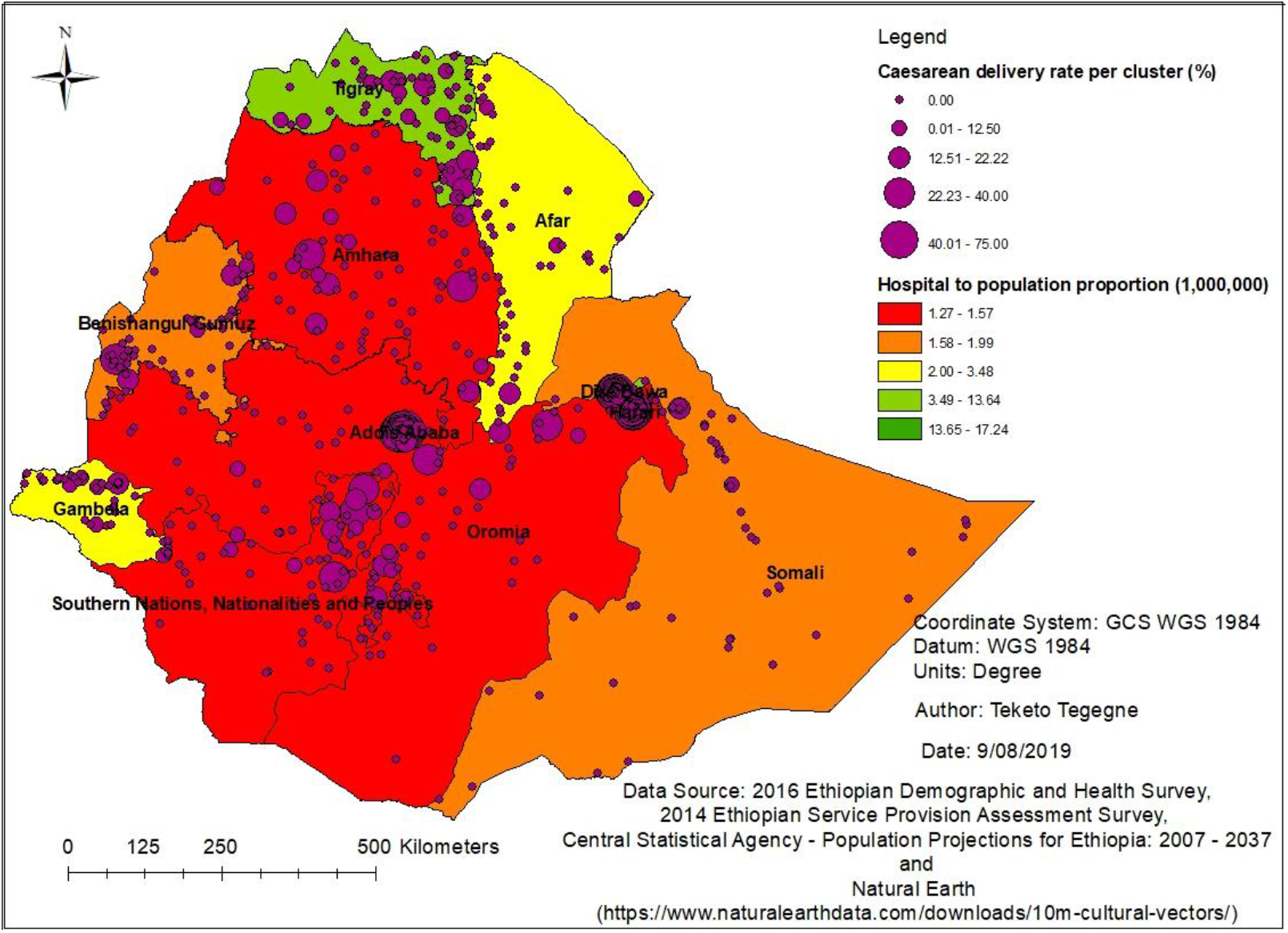
Distribution of hospitals and caesarean delivery rate in Ethiopia, 2016

### Determinants of caesarean delivery use

The calculated EDHS cluster level intra-class correlation coefficient (*ICC*_*c*_) was 15.95%. This indicated that about 16% of variations in using caesarean delivery exist between EDHS clusters. Similarly, the estimated region level ICC (*ICC*_*R*_) indicated that 33.59% of variations in caesarean section rates exist between regions. This indicated that only 50.46% of variability is accounted for by the differing characteristics of the women, or other unmeasured factors. A practically meaningful proportion of variance in caesarean delivery use exists at the EDHS cluster and region levels, providing support for using three-level analytical model.

Women’s age and education, age at first childbirth, household wealth, parity, and ANC visits were significant individual-level caesarean section predictors. A one-year increase in the age of a woman was associated with a 28.13 times increase in the odds of caesarean delivery use. Similarly, a single-year increase in the age of women at first childbirth was associated with a 7.27 times increase in the odds of caesarean delivery. Women who had education were 2.80 times more likely to deliver via caesarean sections compared to those who had no education. A woman from a rich household was 1.80 times more likely to have caesarean delivery relative to a woman from a poor household. Women who had ≥4 ANC visits were 4.67 times more likely to use caesarean delivery compared to those who had no ANC visits. On the other hand, a one-child increase in the number of live births a woman had was associated with a 97% decrease in the odds of caesarean delivery use (Table 1).

**Table 1:** Factors associated with caesarean delivery use among pregnant women in Ethiopia, 2016

| Predictors |  | Adjusted Odds Ratio (AOR) with 95% Credible Interval (CrI) |  |  |  |
| --- | --- | --- | --- | --- | --- |
|  |  | Model 0 | Model 1 | Model 2 | Model 3 |
| **Level-1 predictor variables** |  |  |  |  |  |
| Women's education | No education |  | 1.00 | 1.00 | 1.00 |
|  | Had education |  | **2.85 (1.85, 4.26)** | **2.83 (1.84, 4.17)** | **2.80 (1.83, 4.19)** |
| Wealth | Poor |  | 1.00 | 1.00 | 1.00 |
|  | Rich |  | **2.03 (1.26, 3.17)** | **1.92 (1.17, 3.07)** | **1.80 (1.08, 2.84)** |
| Number of ANC visits | No visits |  | 1.00 | 1.00 | 1.00 |
|  | 1 - 3 visits |  | **3.08 (1.42, 6.22)** | **3.01 (1.40, 6.12)** | **2.91 (1.33, 5.82)** |
|  | ≥4 visits |  | **5.01 (2.40, 9.98)** | **4.91 (2.36, 9.89)** | **4.67 (2.17, 9.43)** |
| Women's age (years) |  |  | **34.05 (6.26, 106.70)** | **31.70 (5.10, 110.30)** | **28.13 (4.53, 98.89)** |
| Parity |  |  | **0.02 (0.01, 0.11)** | **0.03 (0.01, 0.13)** | **0.03 (0.01, 0.16)** |
| Age at 1<sup>st</sup> childbirth (years) |  |  | **5.82 (1.21, 18.13)** | **7.12 (1.30, 22.99)** | **7.27 (1.34, 23.92)** |
| **Level-2 predictor variable** |  |  |  |  |  |
| Distance from caesarean section providing facility (km) |  |  |  |  | **0.12 (0.01, 0.78)** |

ANC = Antenatal Care, km = kilometer, AOR = Adjusted Odds Ratio; CrI = Credible Interval; Model 0 is the null model, a baseline model without any determinant variable; Model 1 is adjusted for individual-level factors; Model 2 is random slope and random intercept model adjusted for individual-level factors; Model 3 is the final model adjusted for individual and cluster-level factors

At the EDHS cluster level (level 2), only one variable was significantly associated with the use of caesarean delivery. A one-kilometer increase in distance to a caesarean section facility was associated with an 88% reduction in the odds of caesarean delivery (AOR = 0.12, 95% CrI: 0.01, 0.78) (Table 1).

Finally, most of the between-EDHS clusters and between-region variances were explained by this model. The proportional change in variance indicated that adding predictors to the empty model explained an increased proportion of variation in caesarean delivery use. The variance estimates between-EDHS clusters decreased from 1.04 in the empty model to 0.05 in the final model. The proportion of the variance of the between-EDHS cluster explained by the final model was 95.19%. Similarly, the variance estimates between-regions decreased from 2.19 in the empty model to 0.14 in the final model. The proportion of the between-region variance explained by the final model was 93.61%. In addition, the empty model showed that 15.95% (*ICC*_*C*_) and 33.5% (*ICC*_*R*_) of the variability in the odds of caesarean delivery use was explained by EDHS-cluster-level and region-level characteristics, respectively. The between-EDHS cluster variability declined over successive models, from 15.95% in the empty model to 1.44% in the final model. Similarly, the between-region variability decreased from 33.59% in the empty model to 4.02% in the final model (Table 2).

**Table 2:** Variations in caesarean delivery use in Ethiopia: random slope and random intercept model

| Random effects (measure of variation for caesarean delivery use) | Model 0 | Model 1 | Model 2 | Model 3 |
| --- | --- | --- | --- | --- |
| Region-level variance | 2.19 | 0.46 | 0.18 | 0.14 |
| Cluster-level variance | 1.04 | 0.11 | 0.07 | 0.05 |
| Variance in women's education (cluster/region) |  |  | 0.07 (0.16) | 0.06 (0.15) |
| Variance in wealth (cluster/region) |  |  | 0.07 (0.17) | 0.06 (0.14) |
| Variance in number of ANC visits (1 – 3 visits) (cluster/region) |  |  | 0.07 (0.19) | 0.06 (0.14) |
| Variance in number of ANC visits ($\geq 4$ visits) (cluster/region) | | | 0.07 (0.17) | 0.06 (0.14) |
| Variance in age (cluster/region) |  |  | 0.06 (0.16) | 0.05 (0.14) |
| Variance in parity (cluster/region) |  |  | 0.06 (0.17) | 0.05 (0.14) |
| Variance in age at 1<sup>st</sup> childbirth (cluster/region) |  |  | 0.06 (0.16) | 0.05 (0.14) |
| ICC<sub>C</sub> (%) | 15.95 | 2.85 | 1.98 | 1.44 |
| ICC<sub>R</sub> (%) | 33.59 | 11.92 | 5.08 | 4.02 |
| Explained variance (PCV<sub>C</sub>) (%) | Reference | 89.42 | 93.27 | 95.19 |
| Explained variance (PCV<sub>R</sub>) (%) | Reference | 79.00 | 91.78 | 93.61 |
| DIC | 1794.45 | 1629.67 | 1628.50 | 1626.78 |

*ICC<sub>C</sub> = Intra-Class Correlation for cluster; ICC<sub>R</sub> = Intra-Class Correlation for region; PCV<sub>C</sub> = Percentage Change in Variance for cluster; PCV<sub>R</sub> = Percentage Change in Variance for region; DIC = Deviance Information Criteria; Model 0 is the null model, a baseline model without any determinant variable; Model 1 is adjusted for individual-level factors; Model 2 is random slope and random intercept model adjusted for individual-level factors; Model 3 is the final model adjusted for individual and cluster-level factors*

### Hotspots of caesarean delivery use

There was evidence of spatial clustering in caesarean delivery use among pregnant women in Ethiopia (Global Moran’s I = 1.15, Z-score = 38.26, P-value < 0.0001). Most of the hotspots were in Addis Ababa, Dire Dawa and Harari, and some parts of the Oromia, Amhara, Somali and SNNP regions. Most of the cold spots were in the Gambela, Benishangul-Gumuz and Tigray regions followed by Amhara and SNNP regions (Fig 4).

**Figure 4:**
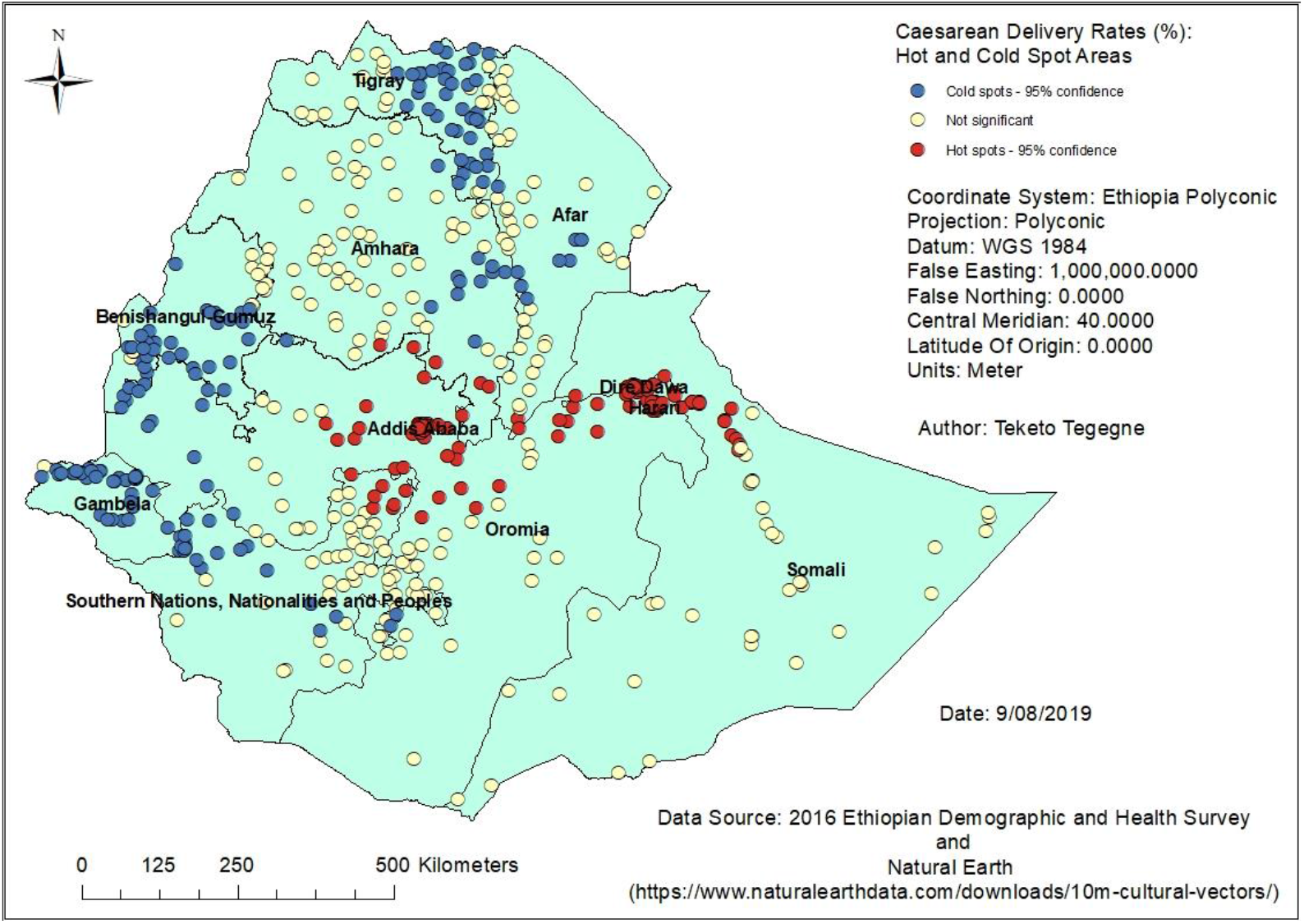
Clusters of high and low caesarean delivery rates in Ethiopia, 2016

### Determinants of spatial variations in caesarean delivery use

In the spatial regression analysis, we found that women’s age and education, parity, ANC visits, age at first childbirth, household wealth, and distance to caesarean delivery facilities were associated with spatial variations in caesarean delivery use. Parity was negatively associated with the spatial variations in caesarean delivery use (Table 3).

**Table 3:** Factors associated with the spatial variations of caesarean delivery use in Ethiopia, 2016

| Variables | Estimate | Standard error | t-value | p-value |
| --- | --- | --- | --- | --- |
| Women's education | 0.54 | 0.22 | 2.42 | 0.02 |
| Rich household | 0.43 | 0.20 | 2.17 | 0.03 |
| 1 – 3 ANC visits | 1.04 | 0.28 | 3.68 | 0.00 |
| ≥4 ANC visits | 1.10 | 0.27 | 4.06 | 0.00 |
| Women's age | 0.57 | 0.26 | 2.22 | 0.03 |
| Age at first childbirth | 0.57 | 0.21 | 2.78 | 0.01 |
| Parity | -0.86 | 0.30 | -2.89 | 0.00 |
| Distance to caesarean delivery facilities | 0.00 | 0.00 | -0.65 | 0.51 |

In the M-SNVC regression model, we found that the relationship between caesarean delivery use and age at first childbirth and ≥4 ANC visits varied across EDHS clusters (Fig 5 & 6). Caesarean delivery use was positively associated with ≥4 ANC visits across Ethiopia (Fig 5). On the other hand, age at first childbirth showed both negative and positive effects on caesarean delivery use across the country. Age at first childbirth showed a strong negative impact on caesarean delivery use in the Tigray and eastern part of Amhara regions and a strong positive impact in the SNNP region (Fig 6). However, the coefficients of the other variables were estimated constant.

**Figure 5:**
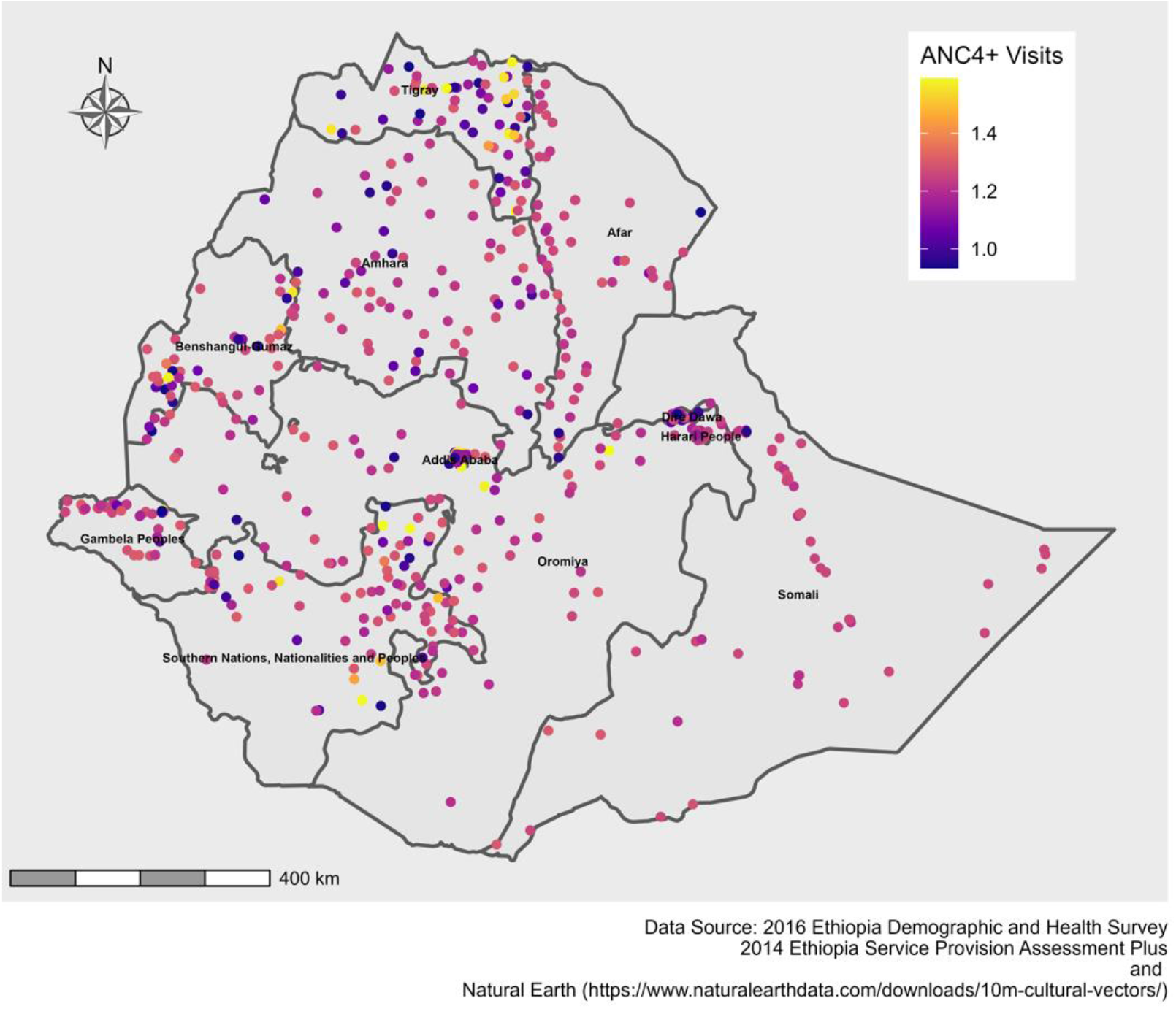
Spatial variations of caesarean delivery use with spatially varying coefficients of ≥4 ANC visits in Ethiopia, 2016

**Figure 6:**
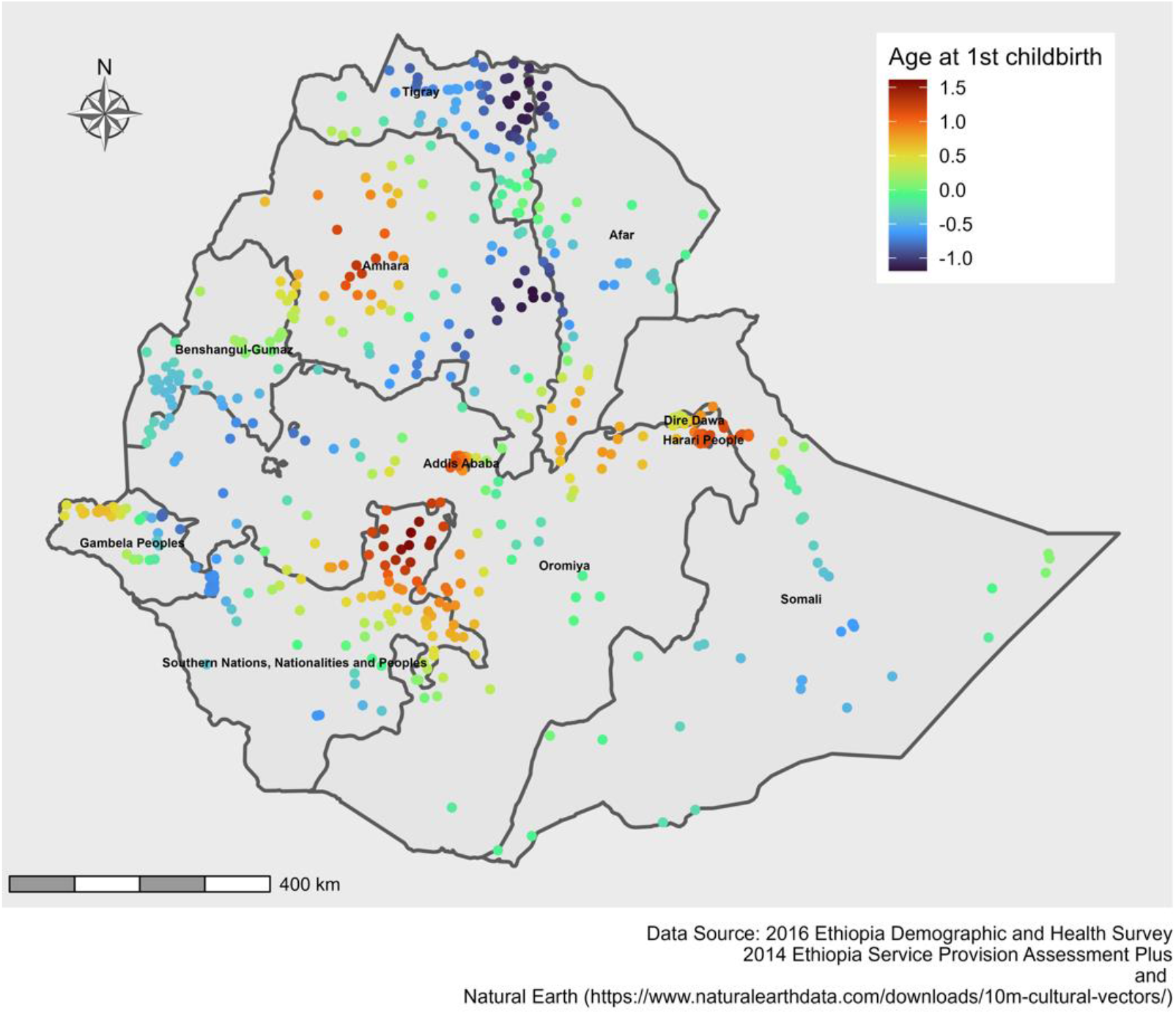
Spatial variations of caesarean delivery use with spatially varying coefficients of age at first childbirth in Ethiopia, 2016

## Discussion

This study provides a comprehensive assessment of unmet needs for caesarean section in Ethiopia by location, service type and demographics. Despite an overall increase in caesarean sections in Ethiopia, there were differences by geographic area, and the rate is lower than the WHO recommendation (36). Furthermore, consistent with previous studies (37-42), caesarean sections in private health facilities were higher than in public hospitals. Caesarean sections for medically indicated reasons can effectively prevent maternal and newborn mortality (43, 44). However, overuse of caesarean sections could increase maternal and perinatal morbidity (45). Private health facilities, which are mainly profit-driven, could do unnecessary caesarean sections to protect themselves from unexpected delivery risks (46). Interventions targeting clinical and non-clinical drivers are required to narrow this gap and avoid unnecessary caesarean sections.

Hotspots of high caesarean section rates were observed in urban centers, i.e., in Addis Ababa, Dire Dawa and the Harari region. This could be due to differences in the distribution of caesarean delivery facilities. Most emergency obstetric care (EmOC) facilities, both public and private, are concentrated in urban areas. These three cities had the highest proportion of hospitals compared to other administrative regions in Ethiopia. Furthermore, in the spatial regression analysis, the geographic variations in caesarean delivery use were explained by women’s age and education, parity, ANC visits, age at first childbirth, household wealth, and distance to caesarean delivery facilities. Spatially varying relations were also observed between caesarean delivery use and ≥4 ANC visits and age at first childbirth. This finding enables informed decision-making for which communities and health facilities need special attention and where government should spend more money.

Women’s use of caesarean sections was influenced by different individual and cluster-level factors. Consistent with previous studies (47-50), a single-year increase in the age of women and age at first childbirth were positively associated with caesarean delivery use. Pregnant women of advanced age are more likely to have pregnancy complications (51-53), comorbidities (54, 55) and higher rates of labour inductions (56). Thus, pregnant women of advanced age should be aware that they might be at increased risk of caesarean section and should prepare in advance. The likelihood of caesarean delivery, consistent with a recent meta-analysis (8), was also positively associated with women’s education. Education could influence women’s decision-making power and delivery mode preference. A systematic review and meta-analysis on women’s caesarean section preference reported that 15.6% of women preferred to give birth via caesarean section (57). It is, therefore, necessary to counsel pregnant women about the potential benefits and risks of caesarean delivery.

Consistent with previous studies (7, 8), women from rich households were more likely to have caesarean delivery compared to women from poor households. Women from rich households could afford expenses related to caesarean delivery and transportation. Furthermore, a single unit increase in the number of live births was significantly associated with lower odds of caesarean section. This finding was consistent with previous studies in Ethiopia (58), India (59), Kenya (60) and Rwanda (6). Multiparas could have faster labour progress compared to nulliparous women (61), supported by the effect of parity on uterine activity study (62). This finding indicates that women in their first pregnancy should get more emphasis during their ANC visits and prepare them to plan skilled delivery care.

Consistent with studies in Rwanda (6), India (42) and Bangladesh (7), the odds of caesarean sections were higher among women with more ANC visits. As per the WHO recommendations, ANC visits are for risk identification, prevention and management of complications, and health education and promotion (63). Women with pregnancy complications identified during ANC visits could have more ANC visits and increased caesarean section rates. In addition, women could prefer caesarean sections if they are uncertain about their fetal condition (64). Thus, the risks and benefits of caesarean and vaginal deliveries should be discussed during ANC visits to avoid unnecessary caesarean sections and birth complications. Further research is needed to investigate why caesarean sections increase along with ANC visits.

Furthermore, a one-kilometer distance increase in caesarean section facility was inversely associated with women’s use of caesarean delivery. Women with medically indicated caesarean sections would not get the service due to limited geographic access to caesarean section facilities (65-68). Thus, limited access to caesarean section facilities could be associated with high maternal and neonatal mortality (9). Despite caesarean service availability and hospital readiness scores did not show significant associations with caesarean delivery, these variables have enormous importance and should not be understated. This non-significant finding could be due to chance or bias, which we could not determine. United Nations recommends that there should be at least five EmOC facilities, including one comprehensive emergency obstetric care facility, per 500,000 population (69), and governments should work to achieve this goal.

The findings of this study should be considered with the following limitations in mind. First, removing EDHS clusters without geographic coordinates could under/overestimate caesarean section rate. Second, misclassification errors due to EDHS coordinates displacement (12) could influence the impact of distance on caesarean delivery use. Third, regarding Bayesian analysis, there is no correct way to choose a prior where we decided to use non-informative priors. Finally, Bayesian analysis often comes with high computational costs (memory and time), especially in models with many parameters and multilevel analysis. Despite these limitations, this study has several strengths. This study used national population and health facility data to identify the demand and supply-side determinants of caesarean delivery use, which most studies investigated separately. Bayesian inferences also solve sample size problems, which is the limitation of classical statistics. We also used spatial analysis to identify geographic variations and associated factors in caesarean delivery use. Investigating geographic variations has a significant role in informed decision-making and monitoring and evaluation purposes.

## Conclusion

The prevalence of caesarean delivery was low in Ethiopia. There were also significant geographic variations across the country, and different individual and community-level factors were associated with caesarean delivery use.

## Data Availability

All data produced in the present work are contained in the manuscript.

## Authors’ contributions

TKT, CC, RS, DL conceptualized the design of the analysis. TKT developed and drafted the manuscript. CC, TG, RS and DL participated in critically revising the intellectual contents of the manuscript. All authors read, provided feedback and approved the final manuscript.

## Acknowledgements

We thank the University of Newcastle, Australia for offering free access to the digital online library to search the electronic databases that were considered for this analysis. We also thank the Measure DHS Program and the Ethiopian Public Health Institute for providing free access to the data sets used for this analysis.

